# Safety and Efficacy of Anti-Fungal agents in Cholangiocarcinoma Patients: A Multi-Center Retrospective Case-Controlled Study

**DOI:** 10.1101/2025.06.16.25329710

**Authors:** Saikat Samadder

## Abstract

**Background:** Comorbid conditions in cancer patients arising from coinfections are one of the main risks of mortality. Recurrent infections are well-known for reducing quality of life in cancer patients. Long-term efficacy of anti-fungal and anti-bacterial agents for drug repositioning is of great importance. In this longitudinal study long term risk and benefits of anti-fungal plus anti-bacterial agents are compared to anti-bacterial agents.

**Methods:** In this multi-center study, patients (n = 3048) diagnosed with cholangiocarcinoma from 1^st^ January 2010 to 1^st^ January 2021 at Severance hospital were analyzed. All patients (n = 983) received standard therapy were included, post propensity score matching (n = 153) patients were included in arm-A treated with anti-bacterial plus anti-fungal agents and (n = 153) in arm-B treated with anti-bacterial agents. Using cox-proportional hazard model, and random forest analyses, efficacy of antimicrobials prescribed during cancer treatment. Post-chemotherapy and post-diagnosis survival in infected were compared among groups. Lastly post infection end-stage leukopenia and leukocytosis events 30 days prior to death were compared.

**Results:** In this study total (n = 306) cholangiocarcinoma patients were included post propensity score matching. There is no significant difference in mean days survived post-cancer diagnosis 712 days (95% CI 605 – 818) arm-A and 615 days (95% CI 536 – 693) survived by arm-B with (*p*-value 0.15). During chemotherapy antibiotic treated arm-A patients survived mean 896 days (95% CI 568 - 1223) compared to arm-B 622 days (95% CI 501 - 742) with *p*-value 0.05. Significant statistical difference in end-stage mortality caused by chemotherapy induced co-infection and leukopenia were found higher in arm-A (14%) compared to arm-B (3%) with (*p*-value 0.05). Post exposure to anti-fungals hazard ratio of fluconazole (n = 113) users compared to non-users (n = 40) of arm-A patients were 1.391 (0.994 – 1.949) with *p*-value 0.07. Hazard ratio of arm-A fluconazole user Vs arm-B teicoplanin treated patients were 1.027 (0.763 – 1.384) with *p*-value 0.8.

**Conclusion:** Patients co-treated with anti-bacterial and anti-fungal agents during chemotherapy survived significantly longer compared to patients treated with anti-bacterial agents alone. Co-treated patients experienced more leukopenia than patients treated with anti-bacterial drugs 8 days prior to death. Mortalities post fluconazole or teicoplanin exposure suggested end-stage fungal or bacterial infections were fatal.

## Introduction

Cholangiocarcinoma along with pancreatic cancer is considered as the fastest growing or aggressive cancer in the world. The prevalence rate of cholangiocarcinoma is highest in China, Thailand, South Korea, and Japan. As per a report of 2020 prevalence rate of cholangiocarcinoma globally was >6 per 100,000 persons per year and mortality 1 to 6 per 100,000 persons per year [1]. Frequent recurrent infections in cancer patients is major concern among health care practitioners due to increased disease burden [2]. Bacterial and fungal coinfections post cancer diagnosis and chemotherapy reduces life expectancy in these patients [3]. Due to availability of wide range of antibiotics for treatment against various types of fungal and bacterial infection has greatly supported the cancer patients [4]. In cancer patients infections caused by *Aspergillus*, *Candida*, *Cryptococcus*, and *Fusarium* species are commonly considered as main cause of aspergillosis, candidiasis, cryptococcosis, and fusariosis respectively [5]. Invasive fungal disease (IFD) caused by these opportunistic fungal strains resulted in prolonged hospitalization of cancer patients [6, 7].

Drugs capable of increasing the quality of life post chemotherapy is in great demand as very few non-chemotherapeutic drugs showed survival benefit and disease free survival in cancer patients [8]. Repositioned drugs such as celecoxib and pemetrexed were found to be efficient against lung cancer patients [9]. Most of the antibiotics like itraconazole, were found to induce tumor cell death in in-vitro studies [10]. Several retrospective longitudinal studies showed few of antibiotics such as itraconazole capable of producing positive impact in cancer patients [11–13]. Identifying an anti-bacterial or anti-fungal drug capable of mitigating the aggressive nature of cholangiocarcinoma or chemotherapy induced mortality could be beneficial.

The first major challenge in drug repositioning of antibiotics is hepatotoxicity arising from prolong usage in cholangiocarcinoma patients with existing liver dysfunction, and antibiotics are not prescribed for prolonged duration [14]. The second challenge arises due to the fact that patients not infected with specific opportunistic microbes included in prospective studies for drug repositioning may not exhibit expected outcome [8]. Immune response against fungal diseases increases IL-17 production capable of inducing tumor apoptosis [15–17]. This may lead to underestimating the contribution of non-severe fungal infection in cancer is the third challenge for estimation of treatment outcome that inhibits IL-17 production. Bacterial infection increases immune upregulation and inflammation resulting in increased TNF-alpha production capable of tumor regression [18].

Despite all the challenges mentioned above in this study the risk and benefit of antibiotics are analyzed. Presently there is no case controlled study available comparing the efficacy of anti-fungal and anti-bacterial drugs under longitudinal study design. A retrospective cohort study including patients with advanced liver disease found fungal infection leading cause of mortality compared to bacterial infections [19]. In another study including pulmonary fungal and bacterial coinfected patients compared to bacterial infected patients for risk identification [20]. In aggressive cholangiocarcinoma very few studies compared the safety of anti-fungal and anti-bacterial drugs, most of the studies demonstrated clinical characteristics or compared efficacy and safety of two anti-fungals [5, 7, 21, 22]. Case controlled study to assess the risk of co-infection is unavailable at present. Here survival benefit are assessed from hazard ratio compared among matched groups post-cancer diagnosis, post-chemotherapy start, and post-antibiotic exposure. Risk of antibiotic failure was identified from deaths occurred 30-days after antibiotic treatment end along with leukopenia and leukocytosis events occurrence.

## Methods

### Study Design

In this multi-center case controlled study, data was obtained from three different branches of Severance hospital, consisting of electronic medical record (EMR) of patients receiving treatment for cholangiocarcinoma. There are (n = 3048) patients diagnosed with cholangiocarcinoma at Severance hospital from 1^st^ January 2010 to 1^st^ January 2021 were identified. All these patients were identified based on searching database with international classification of diseases-10 (ICD-10) codes such as C22, C23 and C24. All necessary prescriptions, initial cancer diagnosis and dates are available for all these patients. Among these patients 2050 were undeceased, insufficient clinical data such as cancer stage, blood tests, patients died within 1^st^ cycle of chemotherapy, patients treated with anti-tuberculosis medications were excluded. After applying first exclusion criteria 998/3048 patients were found deceased until 28^th^ February 2023. Total (983/998) patients were treated with antibiotics prior to death, (n = 15) patients found untreated with antibiotics were further excluded. Total 983 patients were included for propensity score matching, (n = 153) patients were found treated with anti-fungal and anti-bacterial agents included in arm-A and in arm-B (n = 153) patients treated with anti-bacterial medications alone prescribed at any given time point from first hospital visit to death (figure 1). There are patients diagnosed with other primary cancer and received chemotherapy before year 2010, later relapsed to cholangiocarcinoma within 2010-2021. There is no missing data for the included patients.

**Figure 1.**
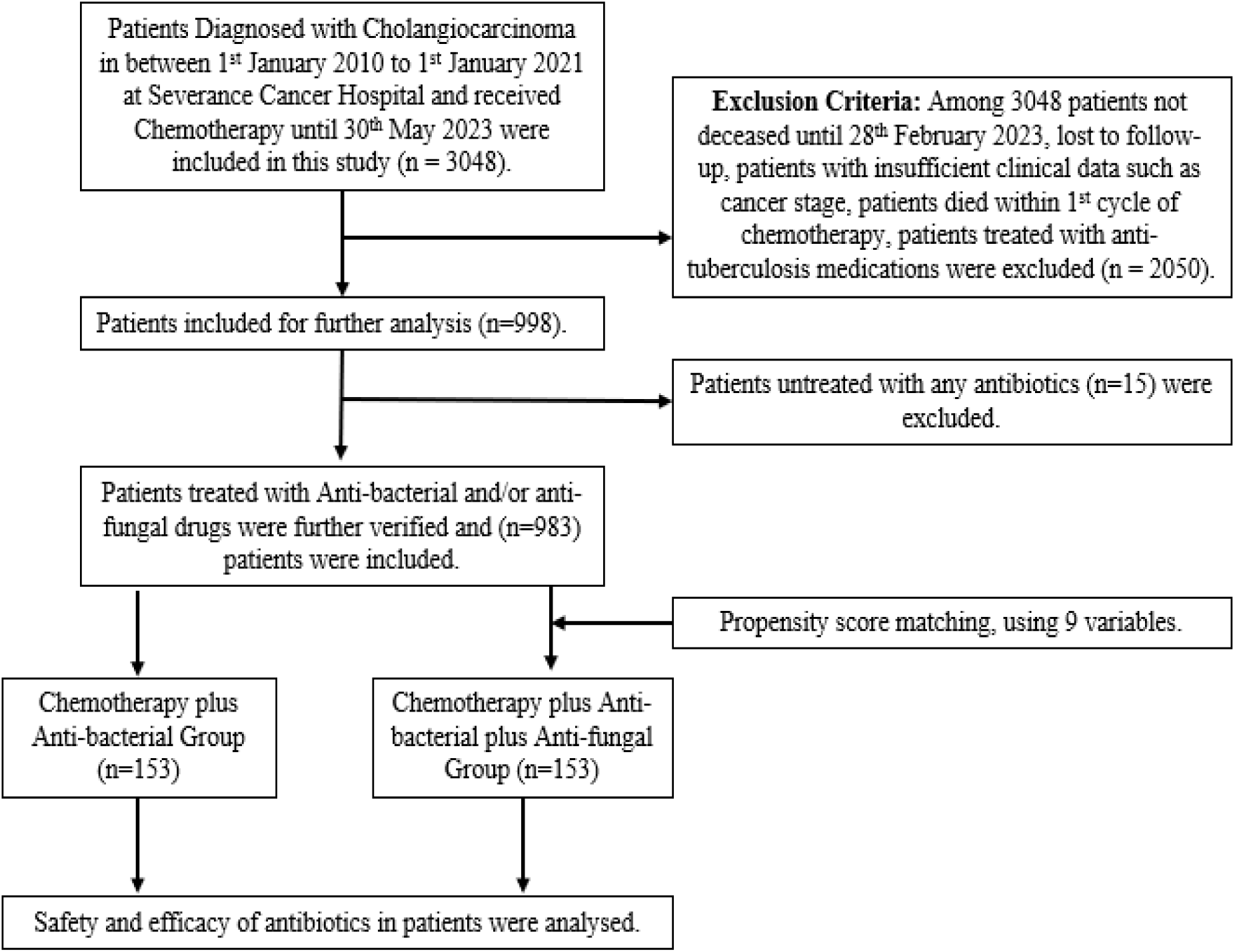
Flowchart for patient Selection.

### Propensity Score Matching

To perform propensity score matching, 10 variables were included during the matching process in RStudio. PSM was matched including gender, age, blood group, cancer location, stage, total chemo dosage, fungal treated/untreated, surgery, and radiotherapy status. In cholangiocarcinoma patients treated with anti-fungal plus anti-bacterial agents were labelled as 1 and matched with anti-fungal non-users labelled as 0; based on algorithm or values matched available from total chemotherapy dosage and 8 other variables [24, 25]. Obtaining standardized difference of between −0.1 to 0.1 post-PSM was considered a sign of balance (**figure 2-3**). PSM using one to many (1:M) was applied in this study allowing to include all patients (n = 153) with triple comorbidity.

**Figure 2:**
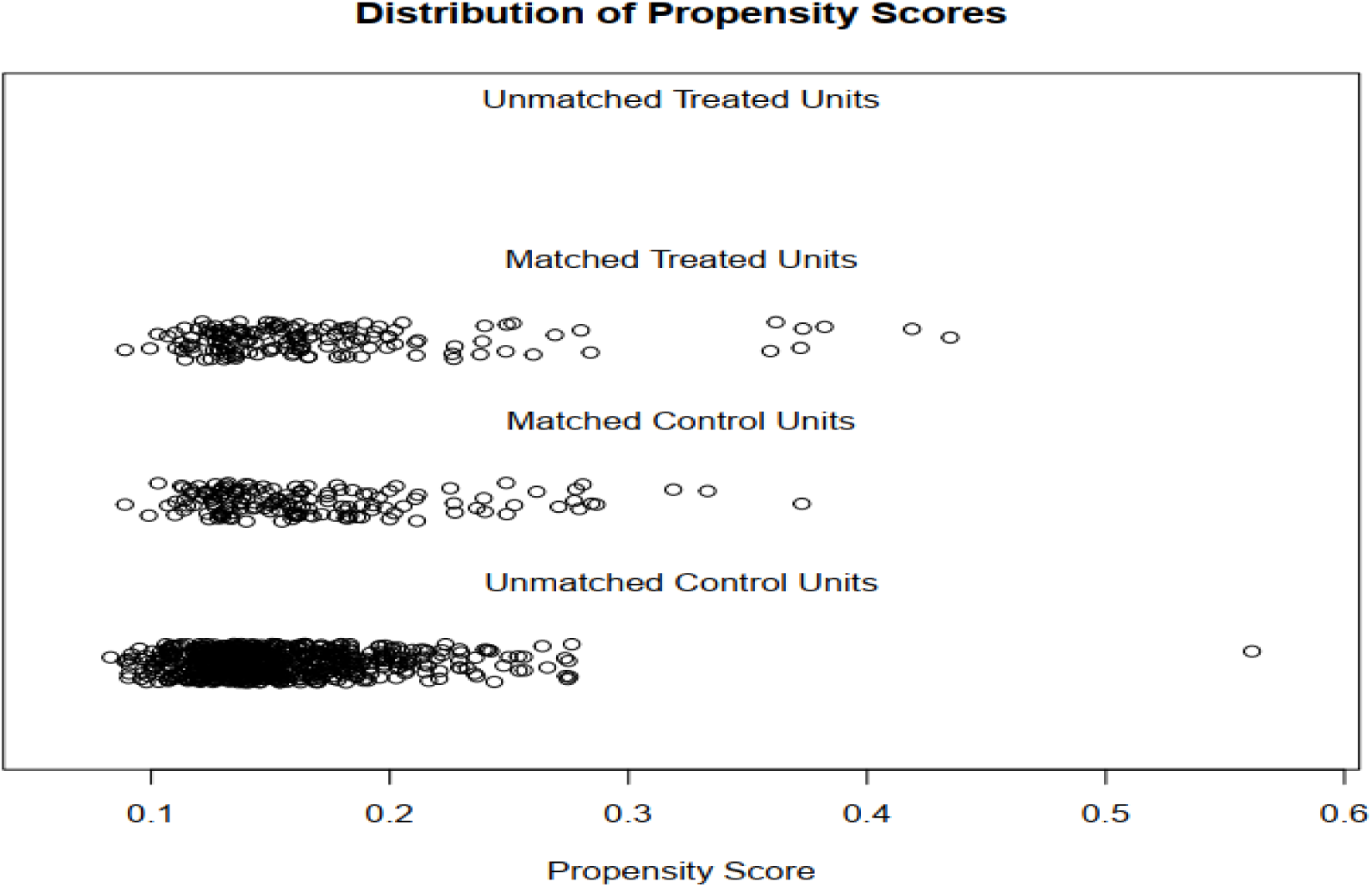
Distribution of propensity score matching.

**Figure 3:**
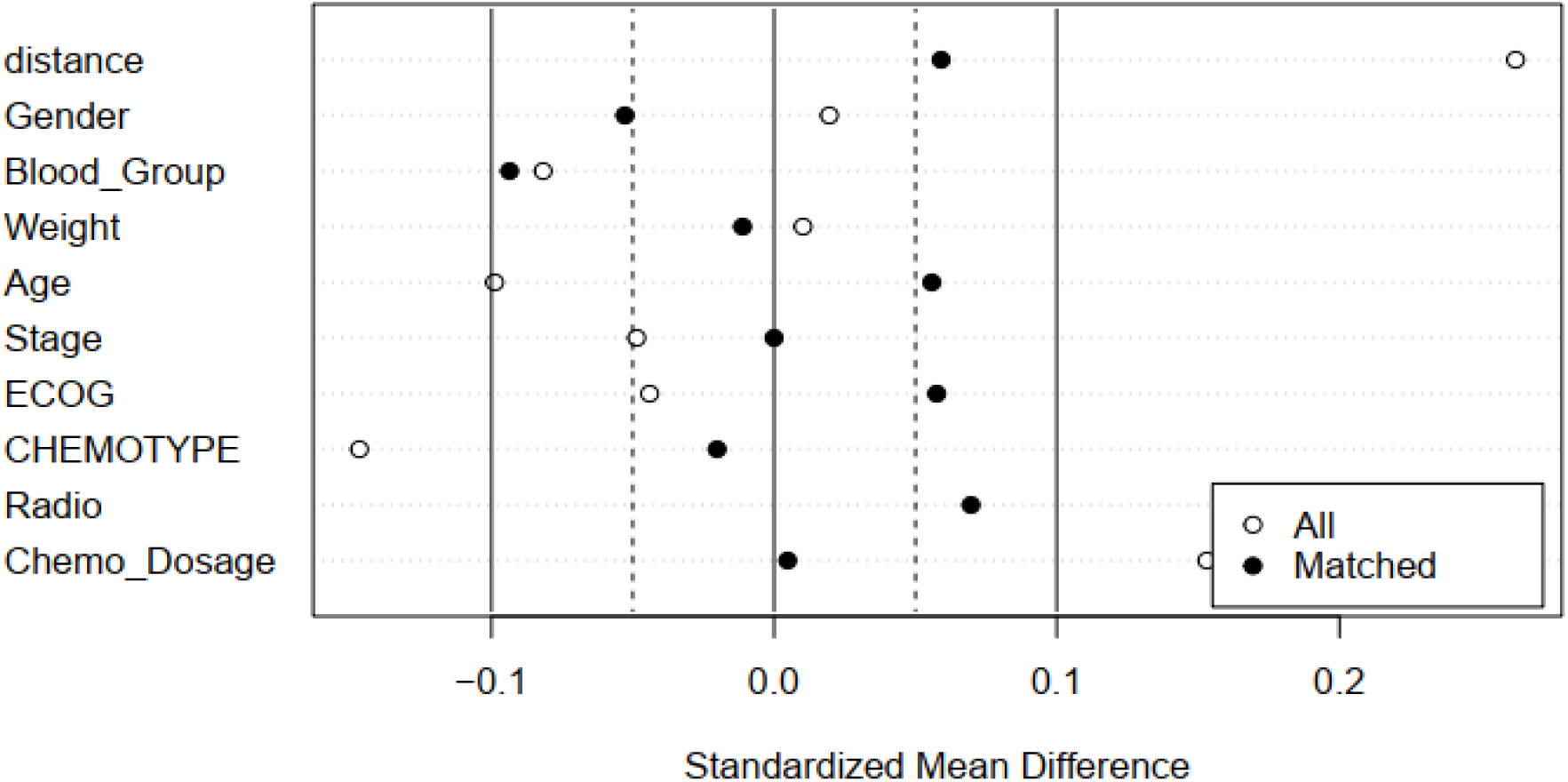
Standardized mean difference **of** nine variables matched and overall samples.

### Antibiotic prescriptions

In this retrospective analysis differential diagnosis (DDx) of any diseased state is a major drawback for retrospective studies evaluating infectious disease. In this study antibiotic prescription dates were considered accurate and allowed to overcome potential disadvantages of DDx. To identify antibiotic prescriptions of all patients (n = 983), all drug prescriptions were extracted from Severance hospital electronic medical record (EMR) database that included in-patient and out-patient prescription records of all patients. All prescriptions available from 1^st^ January 2005 to June 2023 were extracted from database. Generic antibiotics were identified from all types of medicinal products prescribed to (n = 306) cholangiocarcinoma patients and further labelled as anti-fungal or anti-bacterial medications. The generic names obtained from patient identification numbers fetched all available anatomical therapeutic chemical (ATC) codes and prescriptions dates, but this lacked the dose of each generic, limiting scope for dose dependent study.

### Chemotherapy Dosage

To identify chemotherapy dose, volume of drug injected per cycle was divided from the body surface area (BSA). The values of each patient’s BSA were pre-calculated and stored in database with high compliance for cholangiocarcinoma patients. This allowed to acquire and calculate total chemotherapy dosage as mg/m^2^ for all patients. Total dosage for oral formulation was calculated from the standard dose in mg available as per formulation type, multiplied by daily recommended dose, and duration of the treatment. Therefore, the total chemotherapy dosage was total of both mg (oral formulation) and mg/m^2^ (injection) further utilized as a variable for PSM based on total chemo dosage. This enabled to match patients with similar dosage toward chemotherapy in arm-A the average dosage was 20,495 mg/m^2^ and 20,630 mg/m^2^ in arm-B with *p*-value 0.9.

### Infection Induced Death Analysis

Infection induced deaths was calculated from last date of antibiotic prescription to date of death available within EMR database. Patients treated with antibiotics 1 month prior to death was identified and WBC counts of all patients (n = 153) were analyzed for its availability. The last WBC counts available for each deceased patient prior or after one month of last antibiotic treatment was further considered to identify leukopenia or leukocytosis events (figure 4). As average WBC counts for each patient during these 2 months prior to death failed to accurately identify hematological events. There is one patient in Arm-B treated with anti-bacterial agent and last WBC measurement was 8 days prior to infection, or 38 days prior to death. In these 2 months total 4522 WBC counts are available for 245 patients. In arm-A (n = 135) patients 2789 times WBC counts were measured with average of 20.5 (range 75) and in arm-B (n=113) patients WBC were measured 1733 times with average of 15.3 (range 77).

**Figure 4:**
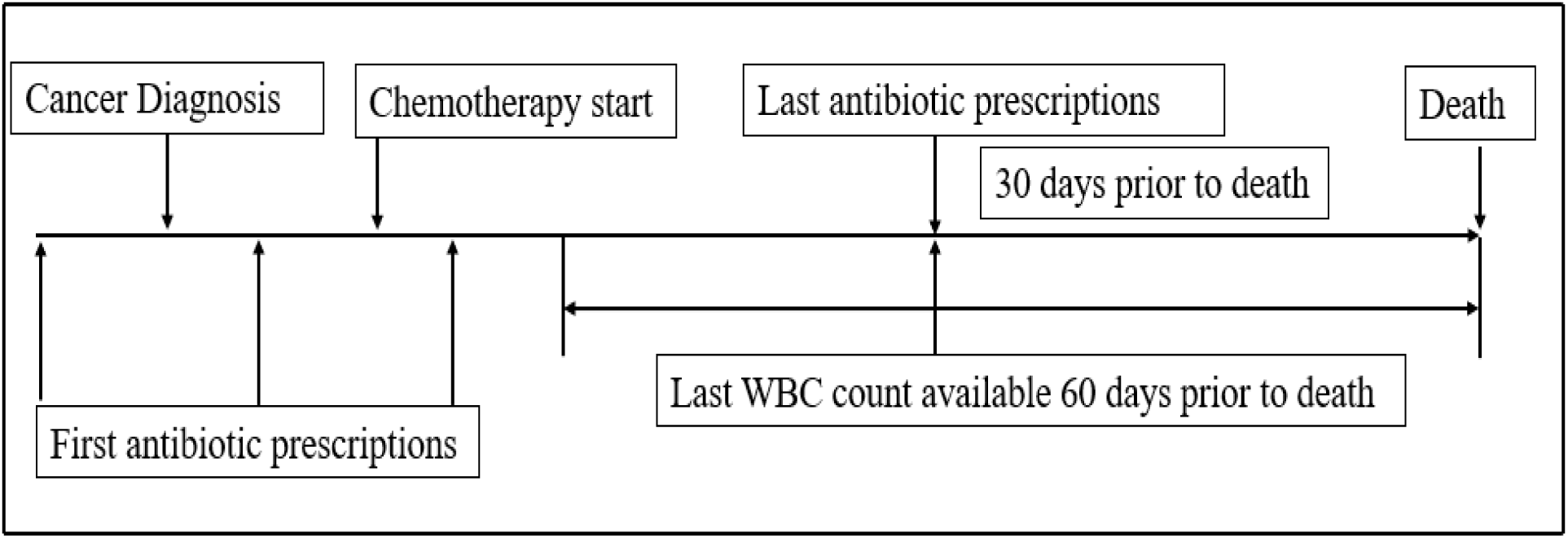
Study design for patient identification and analysis of Infection induced deaths.

### Statistical Analysis

To estimate the safety of antibiotic prescriptions, at first hazard ratio (HR) post-chemotherapy initiation in overall patients, and patients receiving antibiotics during chemotherapy were compared among groups. Safety was further estimated in both the groups and sub-groups based on mortality rates calculated from infection induced death analysis and rate of event occurrence prior to death among groups. To find the HR, days survived post chemotherapy start to death, days survived post cancer diagnosis and post antibiotic start to death was taken into consideration for calculation and comparison among arm-A and arm-B. Using cox-proportional hazard regression analysis mean days survived were compared. One anti-fungal agent (fluconazole), and four anti-bacterial agents (piperacillin, ceftriaxone, teicoplanin, meropenem) were found prescribed to maximum patients of both groups. Efficacy was estimated from 6-months or 1-year hazard ratio and 95% confidence interval (95% CI) calculated from start of prescription to death in anti-fungal and anti-bacterial medications are shown using random forest plot. The baseline characteristics were shown using Pearson’s chi-squared test and welch two sample t-test, time-dependent HR using non-linear regression analysis, PSM, Kaplan-Meier curve lastly forest plot were performed using R (ver. 3.6.1; R Foundation for Statistical Computing, Vienna, Austria). The calculation of 95% confidence interval (95% CI), the *p*-value for HR, mean days survived, and mortality rate was performed online using MedCalc and GIGAcalculator [23, 24].

## Results

### Patient Demographics

After propensity score matching 44.4% females and 55.6% males, mean age of diagnosis was 62.8 years, mean body weight was 60.6 kg, 4 (1.3%) patients at stage-I, 24 (7.8%) patients at stage-II, 76 (24.8%) patients at stage-III, 200 (65.4%) were in stage-IV, and remaining 2 (0.7%) were found at recurred stage. In (n =303) 99% patients ECOG performance status-0 and 3 patients (1%) were found to be ECOG status-1. Surgical interventions were provided specifically for cancer 141 patients (46.1%), and 95 patients (31%) received radiation therapy.

Among 306 deceased patients chemotherapy types (24.5%) adjuvant, (3.9%) neoadjuvant, (71.2%) palliative care, and (0.3%) salvage therapy. The origin of cancer during first diagnosis was primary or secondary cholangiocarcinoma (55.9%), primary other organs (39.2%), unknown and unspecified were (5.2%) patients. Among (n =111) secondary cholangiocarcinoma patients as primary diagnosis includes patients with initial diagnosis with lymph node metastasis. There is no statistical significance between arm-A anti-fungal and anti-bacterial group compared to anti-bacterial arm-B. In overall (n = 306) patients blood test at cancer diagnosis found mean of platelet count (262.2), WBC (7.9), RBC (3.9), bilirubin (3.1), AST (78.8), ALT (90.1), ALP (235.7), albumin (3.8), and CEA (134.8) (figure 5 & 6).

**Figure 5:**
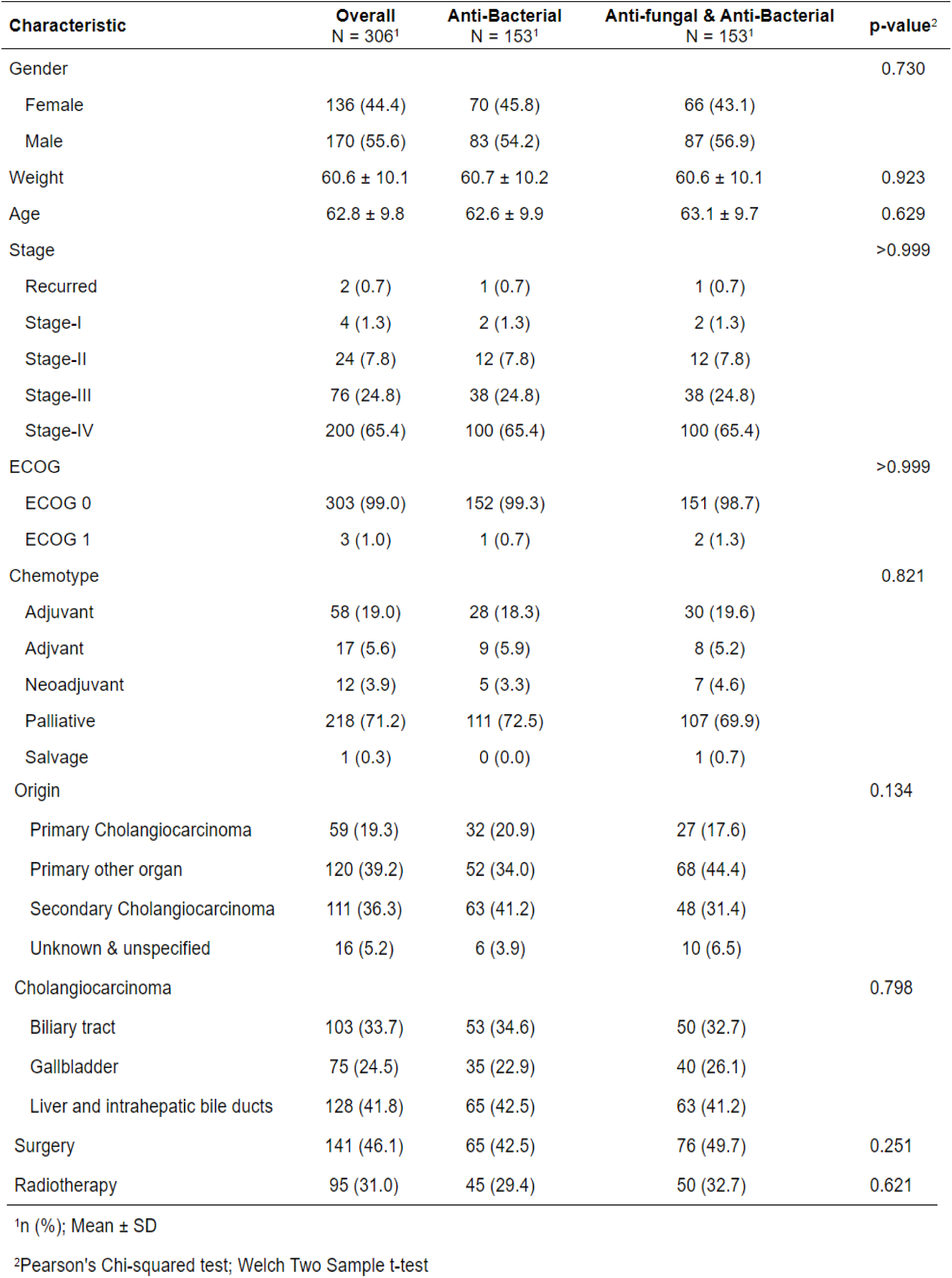
Patient characteristics and baseline.

**Figure 6:**
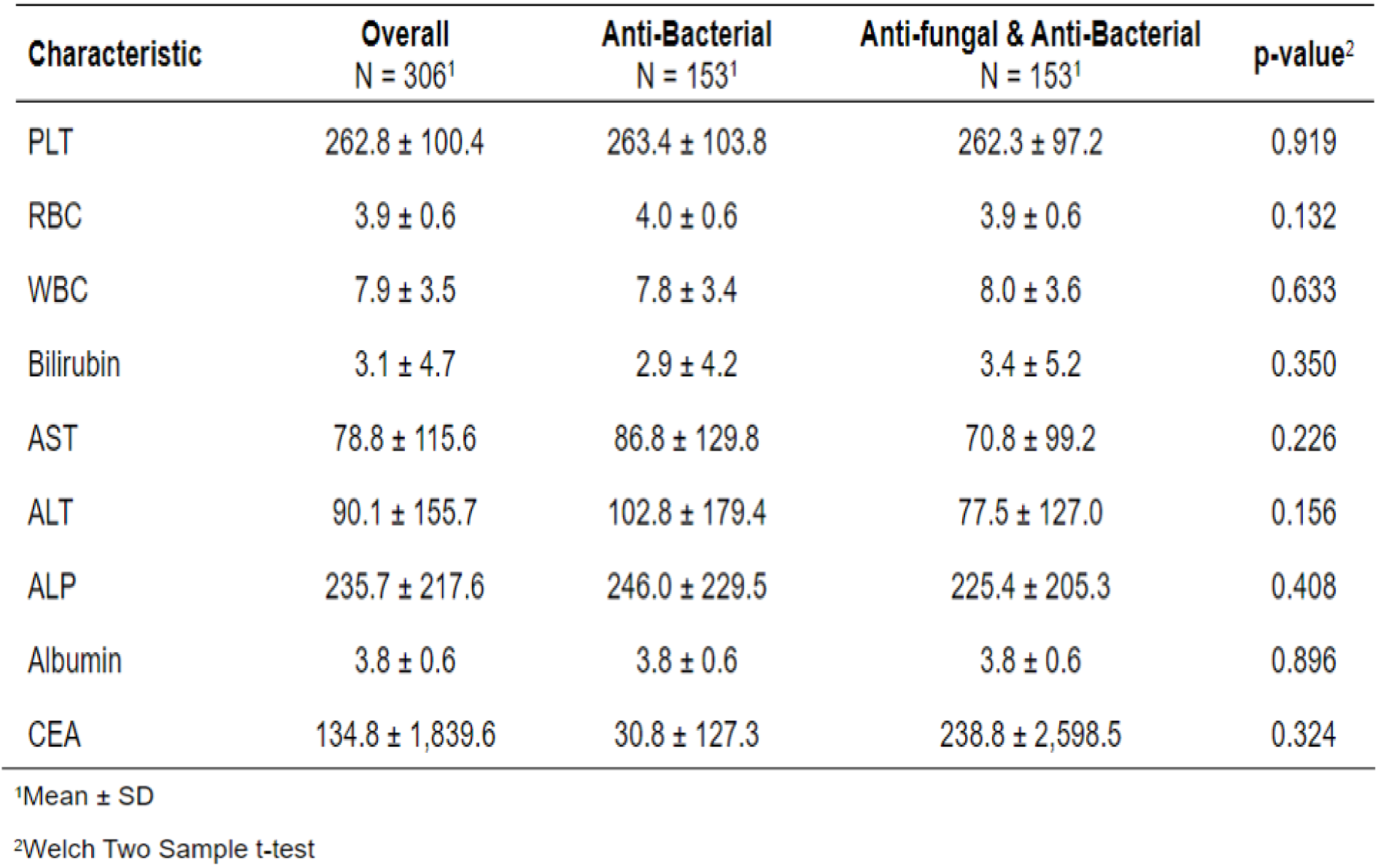
Baseline lab values of blood analyses at diagnosis of cancer for arm-A and arm-B patients.

### Initial Diagnosis of Infection

To analyze the efficacy of antibiotics initial type of infection was identified from antibiotic prescriptions. After initial diagnosis of bacterial infections, 8/153 patients in anti-fungal group (arm-A) were co-treated with anti-fungal and anti-bacterial medications, remaining all patients received anti-fungal drugs post anti-bacterial therapy. Post cancer diagnosis fungal infections occurred in majority of patients post bacterial infection. In anti-fungal group first bacterial infections were detected pre diagnosis of cancer in (n = 51) patients, remaining (n = 102) patients were prescribed anti-bacterial agents initially post cancer diagnosis. In anti-fungal group anti-fungal treatments were prescribed to (n =10) patients initially pre-cancer diagnosis and (n = 143) patients were prescribed anti-fungal treatments post cancer diagnosis. In anti-fungal treated group (n = 114) patients treated with anti-bacterial agents initially prior to chemotherapy initiation, (n = 39) received first treatment with anti-bacterial medicines post chemotherapy start. First anti-fungal agents were prescribed to (n = 18) patients pre-chemotherapy and (n = 135) patients were treated with anti-fungal agents initially post-chemotherapy. In anti-bacterial treatment group (Arm-B) patients (n = 36) were found to be pre-treated with anti-bacterial agents prior to first cancer diagnosis, (n = 117) patients treated with initial anti-bacterial agents post-cancer diagnosis. Anti-bacterial treatment group patients (n = 107) received initial anti-bacterial agents pre-chemotherapy, (n = 46) treated with anti-bacterial medications for the first time post-chemotherapy (Table 1).

**Table 1:**
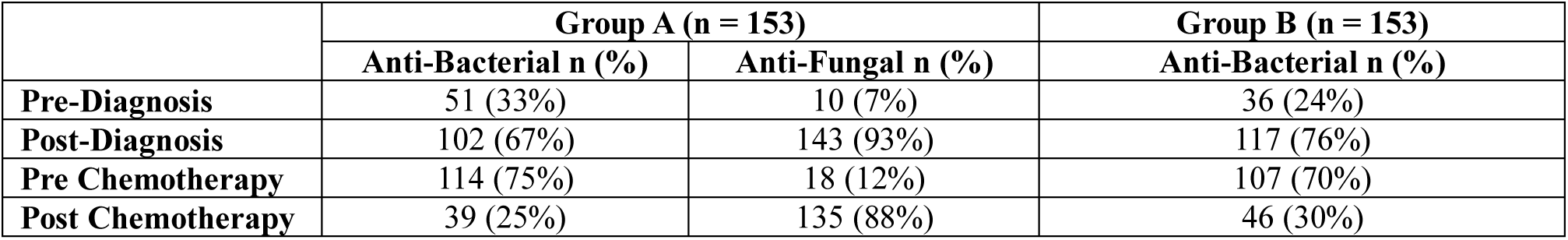
Number of (n) patients treated with anti-bacterial plus anti-fungal agents (Group A) and anti-bacterial agents (Group B) for the first time pre/post-diagnosis of cholangiocarcinoma and pre/post-chemotherapy, respectively.

### Comparison of Overall survival

Anti-microbial agents mean prescription frequency was 140.5 in arm-A compared to arm-B group 66.4 anti-bacterial prescriptions alone with statistical significance of *p*-value <0.001. Majority of bacterial and fungal infections occurred with the onset of cancer or after cancer diagnosis. Fungal infections occurred mostly post chemotherapy initiation (refer Table 1). In arm-A (n = 51) patients prescribed anti-bacterial agents 1396 days (95% CI 935 – 1856) prior to cancer diagnosis compared to arm-B (n = 36) prescribed 452 days (95% CI 160 - 743) with significant statistical difference of 944 days (95% CI 334 – 1553) with *p*-value 0.002. Therefore, due to this difference in initial prescription among groups, post-infection survival largely varied among groups. Consequently 1-year HR post anti-bacterial drug exposure could not be compared accurately. Days survived post initial bacterial infection in arm-A (n = 153) patients 1130 days (95% CI 922 – 1337) compared to arm-b (n = 143) patients 661 (95% CI 548 – 773) *p*-value <0.001. Days survived post fungal infection in arm-A patients was 366 days (95% CI 218 – 513). Days survived post cancer diagnosis in arm-A 713 days (95% CI 605 – 818) compared to arm-B 616 days (95% CI 536 – 693) with *p*-value 0.152 (table 2). Total chemotherapy dosage in arm-A patients was 20,630 mg/m^2^ can in arm-B 20,360 mg/m^2^ without any statistical significance (*p*-value 0.961). Post chemotherapy days survived by arm-A patients 702 days (95% CI 572 – 831) and arm-B 590 days (95% CI 494 – 685) with *p*-value 0.174. Days survived post-chemotherapy is non-significantly different in patients diagnosed with primary or secondary cholangiocarcinoma during first diagnosis arm-A (n = 75) and arm-B (n = 95) 540 days (95% CI 409 – 670) with *p*-value 0.418.

**Table 2:** Days survived post initial infection, post-cancer diagnosis, and post-chemotherapy.

|  | Days survived post initial<br>bacterial infection |  | Days survived post cancer<br>diagnosis |  | Days survived post<br>chemotherapy |  |
| --- | --- | --- | --- | --- | --- | --- |
|  | Arm-A (n = 153) | Arm-B (n = 153) | Arm-A (n = 153) | Arm-B (n = 153) | Arm-A (n = 153) | Arm-B (n = 153) |
| Median days<br>survived (95% CI) | 1130 (922 –<br>1337) | 661 (548 –<br>773) | 713 (605 –<br>818) | 616 (536 –<br>693) | 702 (572 –<br>831) | 590<br>(494 – 685) |
| 1-year Deaths (n) | 50 | 70 | 52 | 65 | 62 | 71 |
| 2-year Deaths (n) | 30 | 30 | 48 | 35 | 42 | 35 |
| 3-5> year Deaths(n) | 44 | 45 | 45 | 49 | 40 | 43 |
| <5-year Deaths (n) | 29 | 8 | 8 | 4 | 9 | 4 |

### Efficacy of Antibiotics during chemotherapy

Comparing the chemotherapy cycles among group suggests arm-A patients received mean cycles of 45.2 and arm-B patients received mean cycles of 58.1 with statistical significance of *p*-value (0.01). Mean duration of chemotherapy in arm-A was 417 days (95% CI 313 – 501) and arm-B was 541 days (95% CI 415 – 667) without statistical difference *p*-value 0.108. Comparison of arm-A (n = 125) to arm-B (n = 111) patients received antibiotics within chemotherapeutic regimen, two years hazard ratio post chemotherapy initiation 0.972 (95% CI 0.784 – 1.204) without statistical significance *p*-value (0.8). Mean days survived post-chemotherapy start by arm-A (n = 125) patients 753 days (95% CI 597 – 908) and arm-B (n = 111) patients 622 days (95% CI 501 – 742). Sub-group analysis of arm-A patients treated with anti-bacterial agents alone (n =76) and anti-bacterial plus anti-fungal agents (n = 49) patients. All these 76 patients received anti-fungal prescriptions before or after chemotherapy. Comparison of arm-A patients (n =49) treated with anti-bacterial plus anti-fungal medications survived 896 days (95% CI 568 – 1223) with statistical significance of *p*-value 0.05. No statistical significance observed (*p*-value 0.684) among arm-A and arm-B patients treated with anti-bacterial medications alone. Time dependent HR post-chemo increased in arm-A (n =125) compared to arm-B (n = 111) without any statistical differences (table 3).

**Table 3:** Median survival (in days) during chemotherapy and antibiotic treatment. Survival calculated post chemotherapy initiation. (*) represents statistical significance between groups *p*-value 0.05. (**^a, b, c, d^**) represents groups and sub-groups. (^b^) represents patients treated with only anti-bacterial drugs during chemotherapy regimen, previously treated with anti-fungal medications.

|  | Arm-A |  |  | Arm-B | *p*-value |
| --- | --- | --- | --- | --- | --- |
|  | All anti-bacterial plus anti-fungal (n = 125)<sup>a</sup> | Anti-bacterial treated patients (n = 76)<sup>b</sup> | Anti-bacterial plus anti-fungal treated patients (n = 49)<sup>c</sup> | All Anti-bacterial patients (n = 111)<sup>d</sup> |  |
| **Median survival (in days) (95% CI)** | 753 (95% CI 597 - 908) | 661 (95% CI 517 - 804) | 896 (95% CI 568 - 1223) | 622 (95% CI 501 - 742) | <sup>a</sup> Vs <sup>d</sup> **0.201**<br><sup>b</sup> Vs <sup>d</sup> **0.684**<br><sup>b</sup> Vs <sup>c</sup> **0.149**<br><sup>c</sup> Vs <sup>d</sup> **0.05\*** |

### Infection Induced Mortality

Infection induced deaths occurred in 135/153 patients 88.2% of arm-A group and in 113/153 patients (73.8%) of arm-B group. Patients died within 8 days after last prescription of antibiotics were considered as severe infection induced deaths and patients died within 8 – 30 days were considered moderate infection. After moderate infections leukopenia and leukocytosis events occurred at similar rate without any statistical significance among groups or sub-groups. There was increased mortality post last anti-fungal plus anti-bacterial drugs prescription and leukopenia events 11% (95% CI 3 – 21) observed in arm-A compared to arm-B (*p*-value 0.005) treated patients with anti-bacterial agents alone prescribed before death (table 4).

**Table 4:**
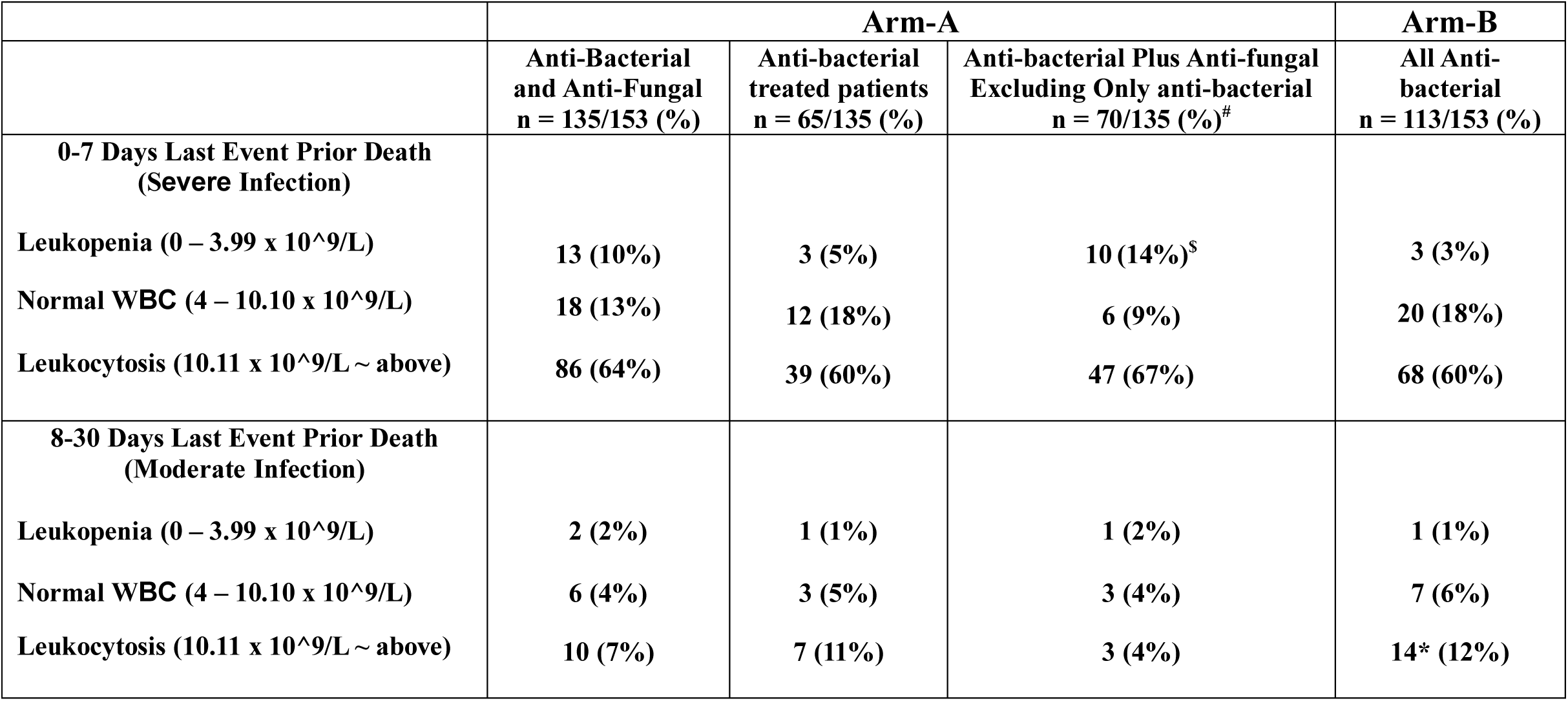
Patients treated with anti-bacterial plus anti-fungal Vs anti-bacterial medications one month prior to death, respectively. Infection induced last events recorded within 8 days (serious) and events after 8 to 30 days (non-serious) prior to death are shown here. (*) represents one patient experienced leukocytosis on 38 days prior to death, 8 days prior to anti-bacterial last treatment.

|  | Arm-A |  |  | Arm-B |
| --- | --- | --- | --- | --- |
|  | Anti-Bacterial<br>and Anti-Fungal<br>n = 135/153 (%) | Anti-bacterial<br>treated patients<br>n = 65/135 (%) | Anti-bacterial Plus Anti-fungal<br>Excluding Only anti-bacterial<br>n = 70/135 (%)<sup>#</sup> | All Anti-bacterial<br>n = 113/153 (%) |
| **0-7 Days Last Event Prior Death**<br>**(Severe Infection)** |  |  |  |  |
| Leukopenia ($0 - 3.99 \times 10^9/L$) | 13 (10%) | 3 (5%) | 10 (14%)<sup>s</sup> | 3 (3%) |
| Normal WBC ($4 - 10.10 \times 10^9/L$) | 18 (13%) | 12 (18%) | 6 (9%) | 20 (18%) |
| Leukocytosis ($10.11 \times 10^9/L \sim$ above) | 86 (64%) | 39 (60%) | 47 (67%) | 68 (60%) |
| **8-30 Days Last Event Prior Death**<br>**(Moderate Infection)** |  |  |  |  |
| Leukopenia ($0 - 3.99 \times 10^9/L$) | 2 (2%) | 1 (1%) | 1 (2%) | 1 (1%) |
| Normal WBC ($4 - 10.10 \times 10^9/L$) | 6 (4%) | 3 (5%) | 3 (4%) | 7 (6%) |
| Leukocytosis ($10.11 \times 10^9/L \sim$ above) | 10 (7%) | 7 (11%) | 3 (4%) | 14* (12%) |

### Efficacy of Antibiotics

Days survived post initial infection was not similar and due to huge difference in days elapsed from first infection to cancer diagnosis in arm-A to arm-B. All the patients in arm-A received anti-bacterial agents prior to anti-fungal agents, because majority of patients received piperacillin in the initial stage for bacterial infections and were prescribed most frequently to both groups being a broad spectrum anti-bacterial. Post drug exposure one year HR of piperacillin treated patients of arm-A compared to arm-B suggests HR 0.8 (0.6 – 1.1) with p-value 0.4, suggesting false positive statistical outcome. Similarly, fluconazole was most frequently prescribed to arm-A patients and comparing 1-yr HR with anti-bacterial alone treatment group, results are false positive statistical outcome (table 5). Since, post anti-fungal exposure mean days survived was 322 days in arm-A and post anti-bacterial treatment survived 661 days in arm-B. Therefore, treated and untreated were further compared, 6 month HR of fluconazole treated patients Vs fluconazole untreated patients of arm-A was 1.78 (95% CI 1.167 – 2.743) with *p*-value 0.02 (figure 7). Six-months HR compared between fluconazole arm-A and teicoplanin arm-B was 1.01 (95% CI 0.72 – 1.41) with *p*-value 0.8 (figure 8). All antibiotic prescription list are presented (table 6-8).

**Figure 7:**
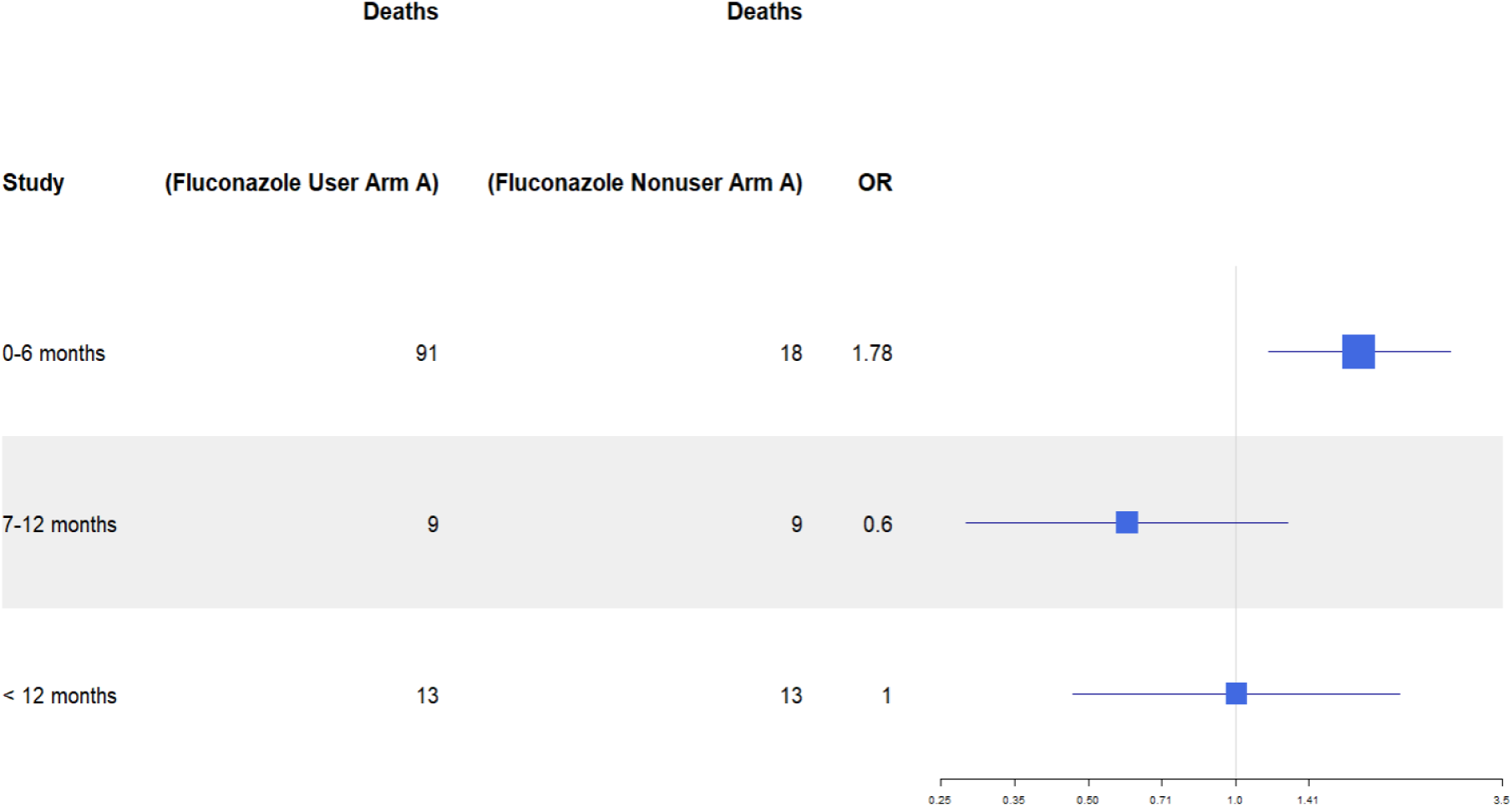
Comparison of post exposure hazard ratio arm-A fluconazole treated with arm-A fluconazole untreated patients.

**Figure 8:**
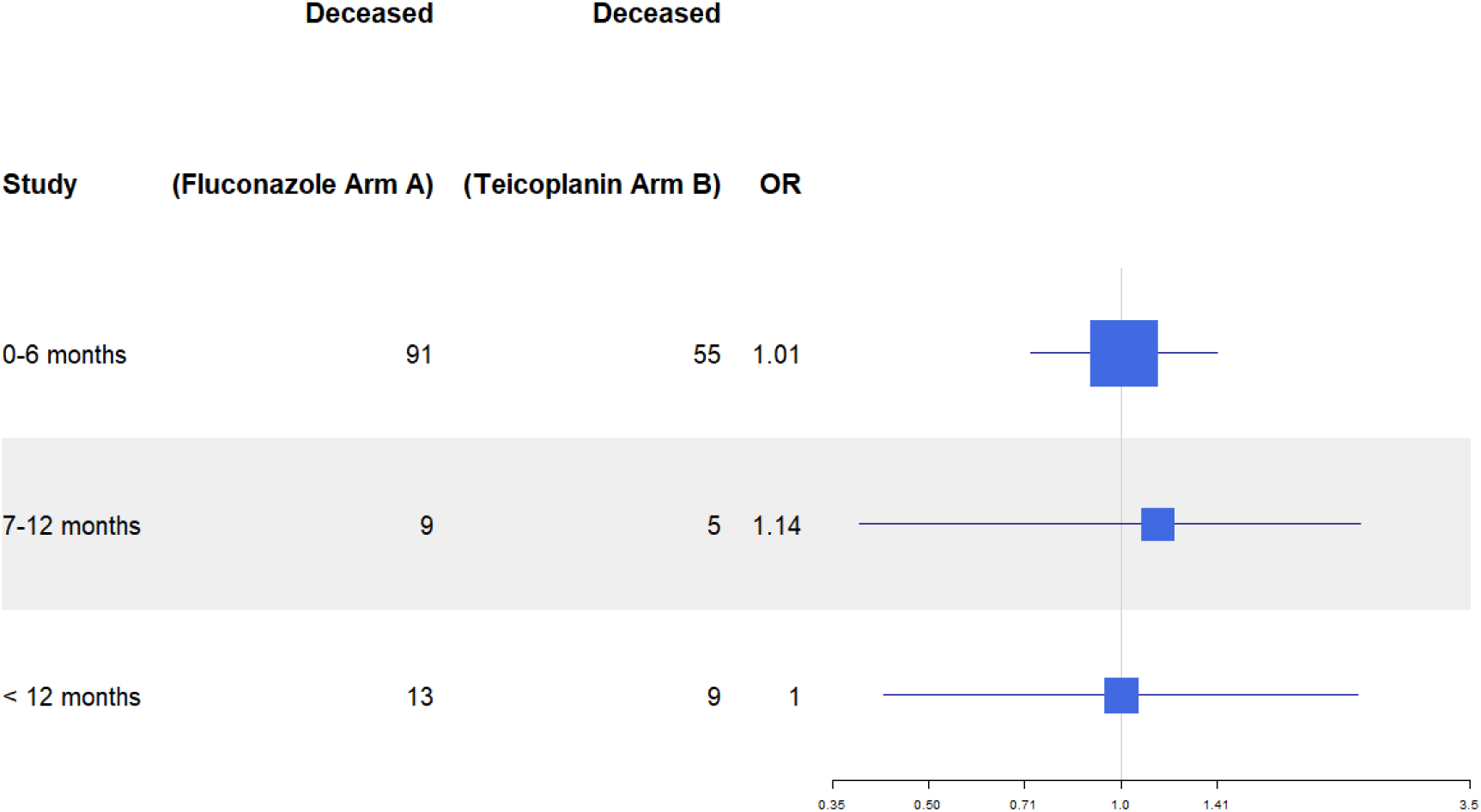
Comparison of post exposure hazard ratio arm-A fluconazole with arm-B teicoplanin exposure.

**Table 5:**
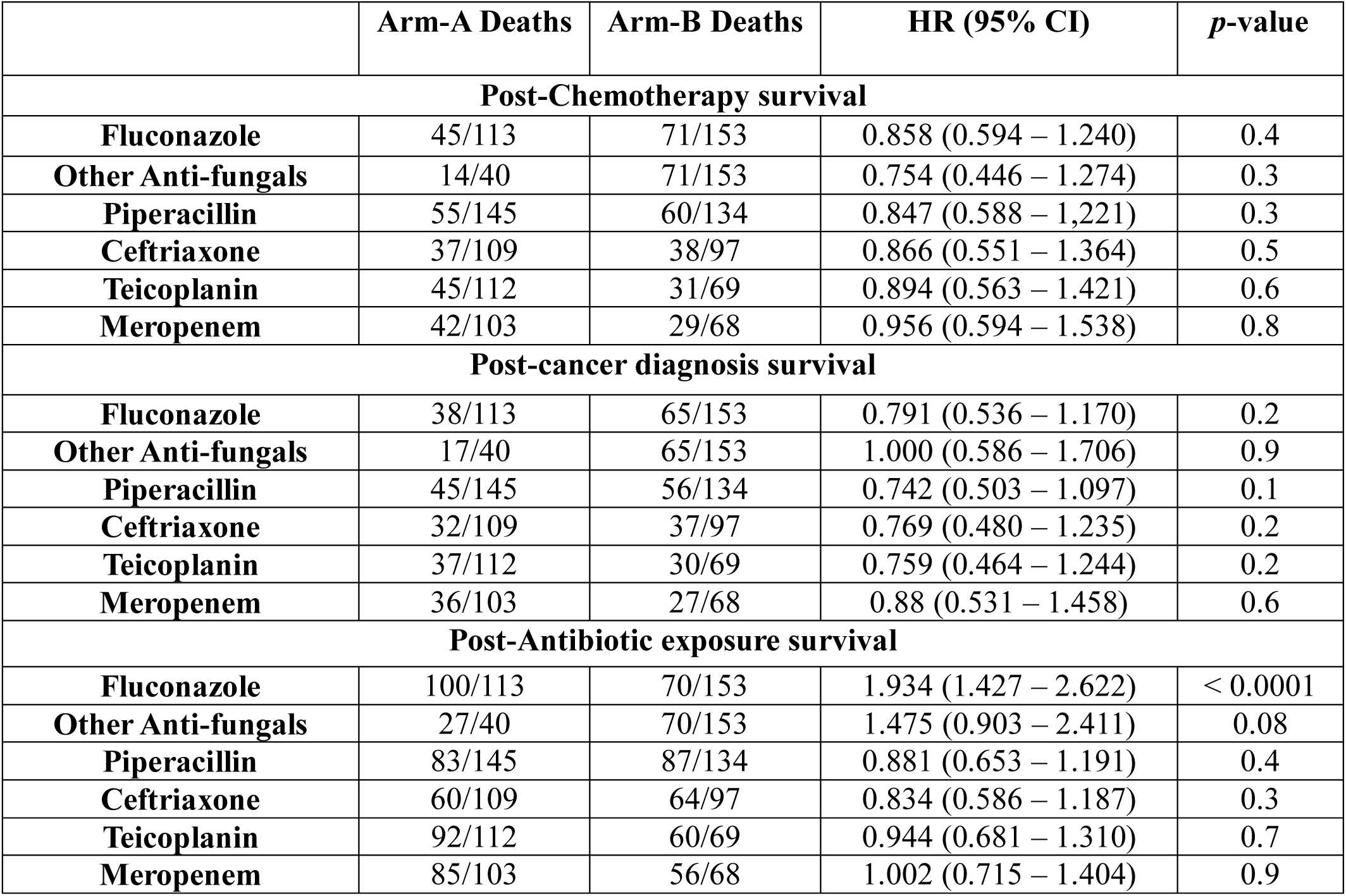
Post antibiotic exposure, post-chemotherapy, and post cancer diagnosis hazard ratio (95% CI) comparison of arm-A Vs arm-B. Although fungal infections occurred mainly post-chemotherapy and bacterial infections occurred post-cancer diagnosis therefore HR of survival post chemotherapy and post cancer diagnosis both are shown. Post antibiotic exposure hazard ratio are false positive.

|  | Arm-A Deaths | Arm-B Deaths | HR (95% CI) | *p*-value |
| --- | --- | --- | --- | --- |
| **Post-Chemotherapy survival** |  |  |  |  |
| **Fluconazole** | 45/113 | 71/153 | 0.858 (0.594 – 1.240) | 0.4 |
| **Other Anti-fungals** | 14/40 | 71/153 | 0.754 (0.446 – 1.274) | 0.3 |
| **Piperacillin** | 55/145 | 60/134 | 0.847 (0.588 – 1,221) | 0.3 |
| **Ceftriaxone** | 37/109 | 38/97 | 0.866 (0.551 – 1.364) | 0.5 |
| **Teicoplanin** | 45/112 | 31/69 | 0.894 (0.563 – 1.421) | 0.6 |
| **Meropenem** | 42/103 | 29/68 | 0.956 (0.594 – 1.538) | 0.8 |
| **Post-cancer diagnosis survival** |  |  |  |  |
| **Fluconazole** | 38/113 | 65/153 | 0.791 (0.536 – 1.170) | 0.2 |
| **Other Anti-fungals** | 17/40 | 65/153 | 1.000 (0.586 – 1.706) | 0.9 |
| **Piperacillin** | 45/145 | 56/134 | 0.742 (0.503 – 1.097) | 0.1 |
| **Ceftriaxone** | 32/109 | 37/97 | 0.769 (0.480 – 1.235) | 0.2 |
| **Teicoplanin** | 37/112 | 30/69 | 0.759 (0.464 – 1.244) | 0.2 |
| **Meropenem** | 36/103 | 27/68 | 0.88 (0.531 – 1.458) | 0.6 |
| **Post-Antibiotic exposure survival** |  |  |  |  |
| **Fluconazole** | 100/113 | 70/153 | 1.934 (1.427 – 2.622) | < 0.0001 |
| **Other Anti-fungals** | 27/40 | 70/153 | 1.475 (0.903 – 2.411) | 0.08 |
| **Piperacillin** | 83/145 | 87/134 | 0.881 (0.653 – 1.191) | 0.4 |
| **Ceftriaxone** | 60/109 | 64/97 | 0.834 (0.586 – 1.187) | 0.3 |
| **Teicoplanin** | 92/112 | 60/69 | 0.944 (0.681 – 1.310) | 0.7 |
| **Meropenem** | 85/103 | 56/68 | 1.002 (0.715 – 1.404) | 0.9 |

**Table 6:**
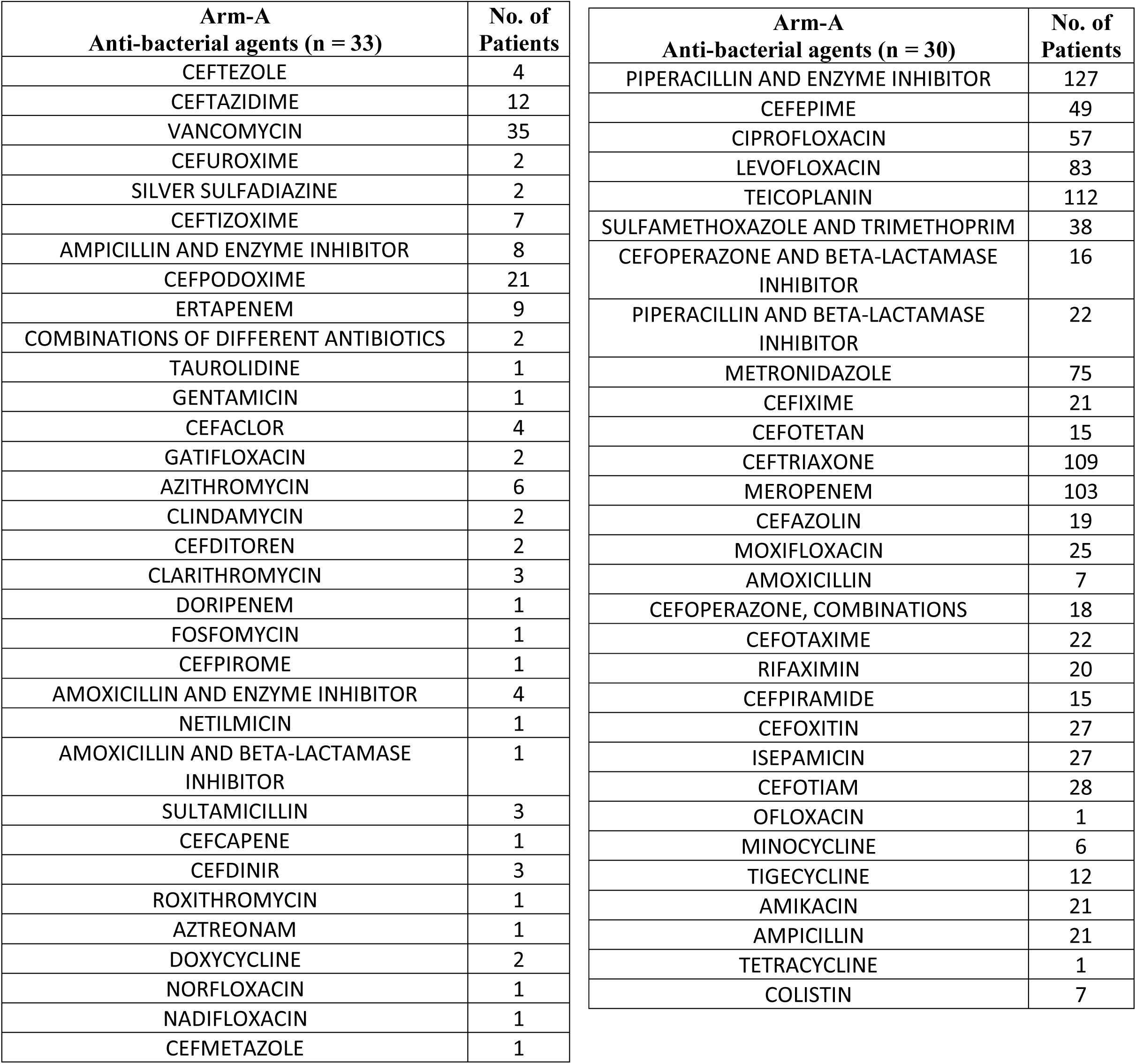
List of anti-bacterial agents prescribed to arm-A of cholangiocarcinoma patients.

**Table 7:** List of anti-fungal agents prescribed to arm-A cholangiocarcinoma patients.

| **Arm-A**<br>**Anti-fungal** | **No. of**<br>**Patients** |
| --- | --- |
| Fluconazole | 113 |
| Isoconazole | 16 |
| Caspofungin | 23 |
| Anidulafungin | 5 |
| Terbinafine | 7 |
| Micafungin | 6 |
| Voriconazole | 8 |
| Amorolfine | 4 |
| Itraconazole | 3 |
| Sertaconazole | 8 |
| Posaconazole | 1 |
| Ketoconazole | 1 |
| Amphotericin b | 8 |

**Table 8:**
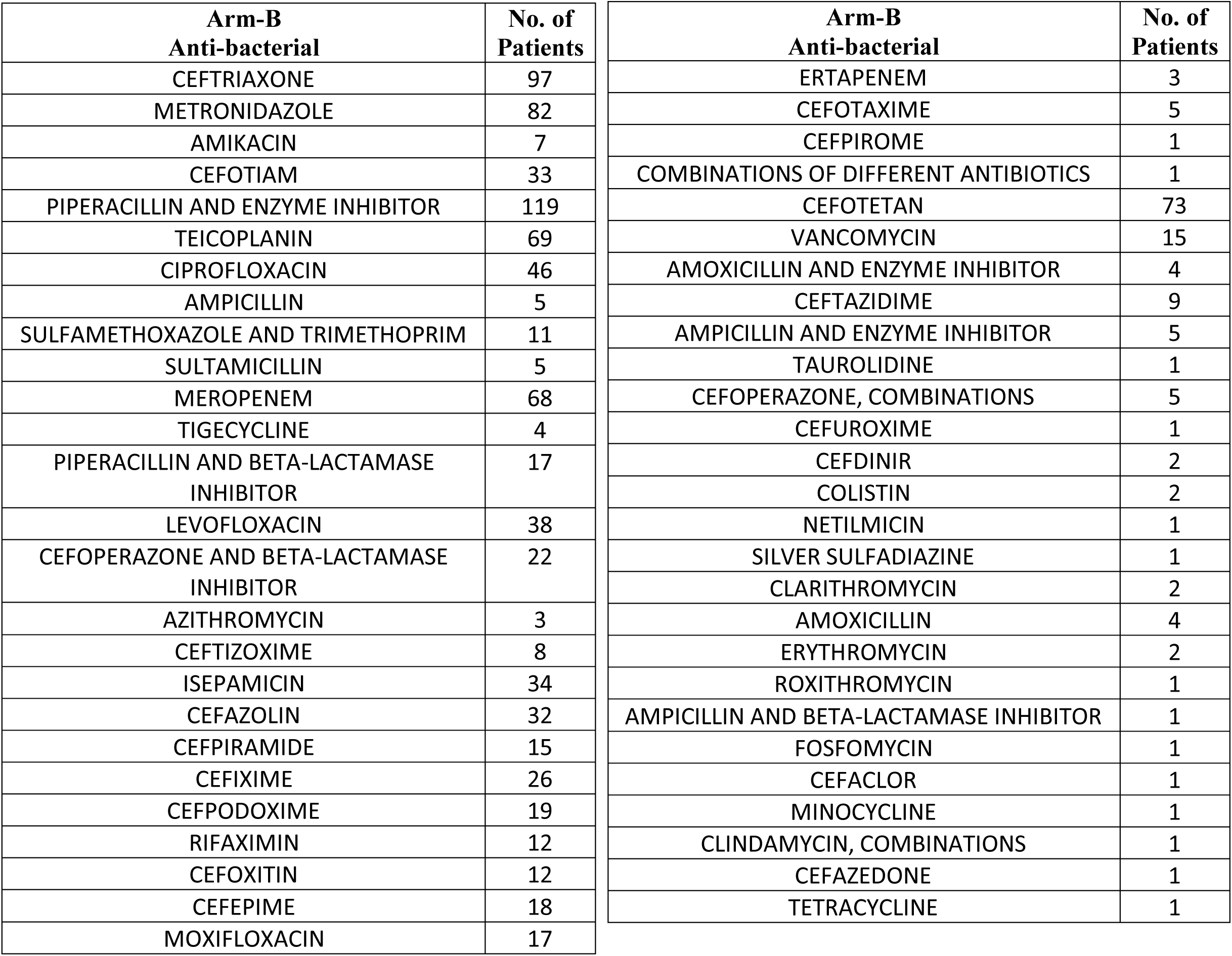
List of anti-bacterial agents prescribed to arm-B of cholangiocarcinoma patients.

## Discussion

Based on antibiotic prescription pattern in this study type of severe infection was found to be candidiasis as fluconazole was most frequently prescribed drugs against candida species [25]. Candidiasis was reported to be most frequently found fungal infection in cholangiocarcinoma patients [3, 26]. There were small proportion of patients prescribed with voriconazole, caspofungin, amphotericin B suggesting these patients were less frequently treated for invasive aspergillosis or invasive candidiasis using these drugs [27–29]. Previously neutropenia were commonly reported in fungal infected patients [30]. In this study post-chemotherapy end leukopenia events were observed at significantly higher rate in fungal and bacterial co-infected patients compared to anti-bacterial treatment group. Neutrophil activation contributes to phagocytosis of fungal pathogens and major a constituent of WBC. Leukocytosis was common in both arms and sub-groups. In anti-bacterial treatment group leukopenia could be caused due to chemotherapy and not related to infection. Here post chemotherapy end survival of leukopenia patients suggested the duration of chemotherapy was related to leukopenia and not related to radiotherapy exposure [31]. Post chemotherapy start and post cancer diagnosis overall survival was not significantly different among groups although triple comorbid condition exists in arm-A. Possibly due to mild initial fungal infection causes increase in IL-17, inhibits tumor growth [16].

Maximum efficacy of anti-fungal drugs were observed during combination therapy of chemotherapy plus anti-bacterial agents. This small sub-population of (n = 49) patients received anti-fungal drugs within chemotherapy regimen remaining (n = 104) patients received anti-fungal either prior to chemotherapy start or after chemotherapy end. Post chemotherapy end fungal infections were rare in overall cancer patients but hazardous [32]. To reduce this risk Tsubamoto et al. in a single-arm retrospective study found itraconazole combined with docetaxel, gemcitabine, and carboplatin to enhance treatment response in bile duct cancer patients. Whereas in this study patients were treated with antifungal drugs at any time during chemotherapy start-to-end based on infection status, here mean days survived post chemotherapy were taken into consideration. In this study (n = 75) patients of arm-A diagnosed with primary and secondary cholangiocarcinoma received combination chemotherapy and antibiotics survived mean 21 months post chemotherapy start compared to itraconazole treated combination chemotherapy patients survived mean 12 months. This survival difference could be mainly due to radiotherapy exposure, in this study 30% and 46% patients received radiotherapy and surgery compared to 7% and 61% patients with bile duct cancer received radiotherapy and surgery with chemotherapy and itraconazole [33–35]. However, during end-stage severe fungal infections most of the (n = 70) patients in arm-A expired within 1 month although there is no significant difference in mortality between groups. Previously, in a retrospective analysis showed that prolonged use of anti-bacterial therapies caused irreversible and severe fungal infection in patients further confirmed in animal model and matches these findings [36]. In this study 208/306 (67%) patients were treated with antibiotics eight days prior to death suggesting chemotherapy induced infection and infection resulted in deaths of these patients.

Main limitations of this study is 1 to many PSM contributing to selection bias. Total 9 variables were considered during PSM could be another limitation, but reducing the number of variables up to 5 does not change significantly days survived by arm-B patients (results not shown). Long term comorbidities such as diabetes or cardiovascular disease were not included at baseline. Events associated with reduced or increased neutrophil, and platelet counts were not presented in this study is another limitation. Contributions of biases from confounding factors such as smoking, drinking, and economic background were not assessed.

Comparison of 6 months HR in fluconazole treated patients with untreated patients of anti-fungal treatment arm suggested fluconazole was not beneficial compared to untreated patients. This indicates that patients were severely infected during fluconazole treatment and previously. Fluconazole were usually prescribed against end-stage severe fungal disease and to bacterial coinfected patients. Further comparison of fluconazole with teicoplanin suggested hazards from fluconazole was not significantly different from teicoplanin treatment. This suggests both were used during severe infections. Teicoplanin is usually prescribed to patients with gram-positive bacterial infection, such as methicillin-resistant *Staphylococcus aureus* (MRSA) [37]. Teicoplanin and fluconazole were reported as hepatotoxic drug both efficient to manage designated infections better than other similar antibiotics such as isoniazid or vancomycin, infections during end-stage of cancer may not be beneficial by these drugs [38, 39]. This study sufficiently evaluated that invasive fungal infection and drug resistant bacterial infections are both equally severe and majorly contributes in mortality of cholangiocarcinoma patients within six months to one year post antibiotic exposure. This finding encourages to repurpose anti-fungal drug more potent and less hepatotoxic in nature against invasive candidiasis pre-diagnosed with cholangiocarcinoma.

Several studies suggested that hepatotoxicity caused by itraconazole is higher than fluconazole in cancer patients, fluconazole was suggested against invasive fungal disease found less beneficial for leukemia patients [40, 41]. To overcome toxicities induced by fluconazole it is suggested here to replace fluconazole with ibrexafungerp for cholangiocarcinoma patients with invasive candidiasis [42, 43]. Presently, ibrexafungerp is indicated for vulvovaginal candidiasis and off-label prescription for IFD was found beneficial to patients due to low toxicity profile [44]. Identification of potent and safe anti-fungal and anti-bacterial for cholangiocarcinoma patients undergoing long-term chemotherapy would be effective against recurrent fungal infections. Future clinical trials on ibrexafungerp against IFD in cholangiocarcinoma patients might substantially reduce mortality and disease burden.

## Supporting information

STROBE checklist

## Acknowledgement

The author would like to thank professor Kyungsoo park (Department of pharmacology, Yonsei University) for providing access to severance hospital database for obtaining patient’s electronic health records.

## Ethics Statement

All relevant ethical guidelines have been followed, and any necessary IRB and/or ethics committee approval was obtained. The studies involving human participants were waived by the Institutional review board (IRB) of Yonsei University because of absence of risk to study participants. Written informed consent form for participation was not required for this study in accordance with the national legislation and the institutional requirements.

## Data availability

Data will be made available post-approval from Yonsei University IRB.

